# Predictive model for real-world performance of COVID-19 antigen tests based on laboratory evaluation

**DOI:** 10.1101/2024.10.21.24315762

**Authors:** Miguel Bosch, Dawlyn Garcia, Lindsey Rudtner, Nol Salcedo, Raul Colmenares, Sina Hoche, Jose G Arocha, Daniela Hall, Adriana Moreno, Irene Bosch

## Abstract

Controlling spread of disease due to infectious agents require a quick response from public health sector. In the ongoing COVID-19 pandemic, the use of antigen tests has shown to be an excellent tool to inform authorities and mitigate the spread of the disease. In this communication we demonstrated how performance of an antigen test -- as a diagnostic *in vitro* device -- can be properly validated using quantitative laboratory experimentation and self-testing data from a clinical study. We also show how clinical performance of an antigen test can be predicted using mathematical modeling. The proposed appraisal methodology of antigen test performance under real-world conditions could be a useful tool to inform regulatory decision making. This approach allows to standardize, democratize, and speed up the process of validation, analysis, and comparison of antigen rapid tests, and thus to help developing effective public health response strategies.

## Introduction

Quantifying the performance of antigen tests for regulatory science, commonly requires the calculation of test performance statistics (e.g. sensitivity) of an antigen test’s binary assessments with reference to the qRT-PCR gold standard. These statistics are based on human clinical samples, requiring paired qRT-PCR testing to generate the validation. Because the antigen test performance is closely related with the sample viral load [1-3] these statistics are dependent on the viral load distribution within the trial dataset. Hence, the regulatory process commonly requires testing under conditions related to the sample size and viral load distribution in qRT-PCR spectrum of low, medium, and high virus content. As the corresponding data involves testing infected subjects, collecting this data requires formal clinical study execution. These processes take typically several months to accomplish.

Rapid tests such as lateral flow assays (LFA) used for antigen detection can be powerful tools for transmission control of viruses such as SARS CoV2. Thus, fast deployment of rapid tests without compromising quality assurance in the evaluation process is key for outbreak control and pandemic preparedness. We have developed a methodology for the quantitative evaluation of COVID-19 antigen tests (ATs) based on laboratory measurements of the test’s band signal intensity and binary naked-eye user assessments. In both cases, we characterized the test performance according to sample concentration of the target recombinant protein, and with heat inactivated virus as well as biologically active virus, and also using real human samples collected under a prospective clinical protocol. Including the statistical characterization of the user’s population LoD in the signal intensity domain, we develop a predictive model for the probability of positive agreement in real-world conditions.

Our method involves: (1) characterizing the AT signal intensity with protein and inactivated virus dilutions, (2) calibrating the qRT-PCR cycles with virus dilutions, (3) characterizing the signal intensity LoD of the user population for the AT, and (4) predicting real-world probability of positive agreement signal response of the AT. Our methods have the advantage of being formulated on continuous variable analysis and probability models, instead of plain discrete analysis and sample statics.

We demonstrate our analyses and compare the predicted probability of positive agreement against real-world data collected from an IRB-approved study that utilizes frequent antigen testing to monitor COVID-19 in an underserved population (Advarra Protocol Number: Pro00059157). Participants consisted of individuals from vulnerable populations in low-income and assisted-living facilities located in Chelsea, MA [4]. Recruitment sites include State-regulated, independent, senior living facilities in Chelsea, MA, and other residents of the city of Chelsea. In the study, participants were provided with antigen tests to self-test routinely for COVID-19 at home or in community centers, uploading the test results and photos to the project informatic platform. We obtained confirmatory qRT-PCR data for all positive results detected by antigen test from an independent CLIA laboratory, including cycle threshold values, and for a random number of negative results.

We describe the quantitative analysis of the AT for signal intensity and naked-eye binary data, the characterization of the user’s LoD, the calibration of the qRT-PCR, and fully describe the formulation of the predictive model. In the result section, we illustrate the application of the described methodology with the characterization of an antigen test in the common cassette device presentation, and compare the predicted result with the real-world probability of positive agreement.

## Methods

The present methodology combines: (1) a quantitative evaluation of the test signal response to concentrations of target protein and inactive virus or active virus, (2) a statistical characterization of the LoD using observer’s visual acuity of test band, (3) a calibration of a gold standard method (e.g. qRT-PCR cycles) against virus concentration. We describe these quantitative methods and unfold a Bayesian based predictive model to describe the real-world performance of the antigen test, quantified by the probability of positive agreement as function of viral-load variables (like qRT-PCR cycles). Figure 1 describes the different components of information involved in the predictive model.

**Figure 1.**
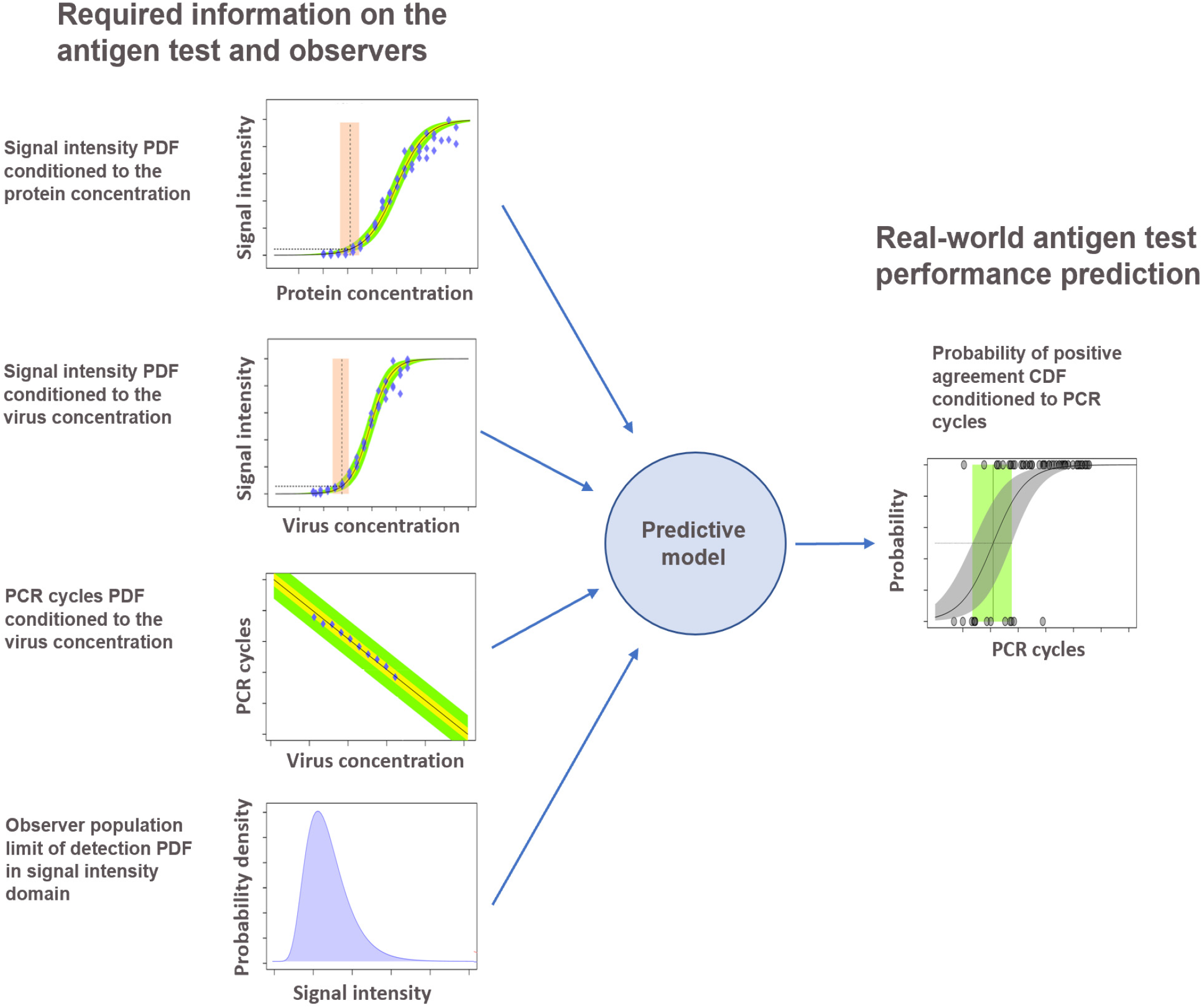
Schematics of the inference model. Input components are based on the laboratory calibration of the antigen test signal intensity against protein and virus concentration, qRT-PCR cycles against virus concentration and the limit of detection probability density function (PDF) of the observer population’s visual assessment of the devices. The result of the model is the performance in ordinary user conditions (*real-world*) of the antigen tests, quantified as the probability of agreement function conditioned by the qRT-PCR value.

### Antigen test signal response characterization

In a common AT device (e.g. cassette or strip) we measure the signal intensity of the test band, white background and control band captured digitally by a photograph using a portable cell phone camera. We process this image and evaluate the average pixel intensity in various areas of the nitrocellulose strip. Measured in this way, the signal intensity is a continuous variable independent of the observer. There are various ways to assess antigen test band signal intensity [5-6]. We calculate the signal intensity subtracting the white background and test band average pixel brightness, and normalizing by the largest signal intensity present in the dataset.

We analyze the signal intensity response of an AT conditioned to the target, calculating the signal intensity across a dilution series of the recombinant protein. To fit these data, we use the Langmuir-Freundlich [7-8] absorption model,

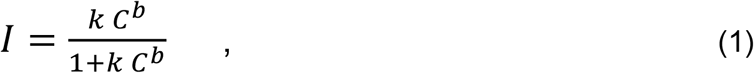

with *I* being the normalized signal intensity, *C* the concentration, *k* the adsorption equilibrium constant and *b* an empirical exponent, close to one. The model parameters to estimate by fitting the normalized intensity data correspond to the adsorption constant, *k*, and the exponent, *b*. We use a Bayesian regression solved with Monte Carlo sampling, which provides description of the model uncertainties. We follow a similar procedure to characterize the relationship of the normalized signal intensity with other variables related to the recombinant protein concentration (e.g. inactive or active virus concentrations).

### Probability of agreement function in naked-eye assessment

Common use of ATs involves human naked-eye interpretation of the result. The outcome of each assessment is a binary variable, either positive or negative (1 for positive and 0 for negative for mathematical analysis). The observer’s visual acuity and ability, understanding the instruction of use would influence the results.

Common measures of the test performance are well-known classification statistics, like the sensitivity. These statistics have a known disadvantage in accurately describing the antigen test performance: they are strongly dependent of the viral concentration distribution of the tested samples. (e.g. the sensitivity improves with a larger representation of strong concentrations).

For an accurate description of the naked-eye performance of the test, we estimate the probability of a positive agreement (PPA) as *function* of the viral concentration, or other viral concentration related variable like the qRT-PCR cycles [9]. We model the PPA with a logistic function,

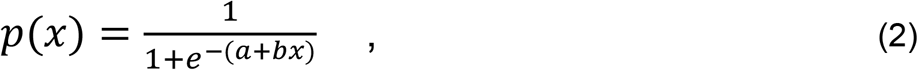

with *x* being the viral load related variable and *p*(*x*) being the probability of positive agreement function. The model parameters to estimate fitting the binary naked-eye data are the *intercept, a*, and the *slope, b*. Description of the of the PPA function in commonly done against the qRT-PCR cycles; similarly, we apply this method for other viral-load related variables (like concentration and normalized signal intensity).

### Predictive model for the probability of positive agreement

The formulation of the model follows a probabilistic approach, meaning that variables and relationships across the model are randomized and described by probability densities functions (PDFs). We define random variables used in this formulation. A group of continuous positive variables that are related with the viral load: the recombinant protein concentration, *x*_prot_, the virus concentration, *x*_vir_, the test normalized signal intensity, *x*_int_, the observer LoD normalized signal intensity, *x*_lod_, and the qRT-PCR cycles, *x*_cycle_. In addition, we have the binary agreement variable, *A*, which indicates the observer assessment of the test outcome, with values 0 (for negative) and 1 (for positive).

For the purpose of this analysis, the limit of detection (LoD) does not represent an exact value. We analyze the LoD associated to a group of observers (i.e. certain population), or for a single observer the LoD, which depends of different environmental circumstances (e.g. illumination, visual context, etc.), and individual abilities. Hence, we consider the LoD as a random variable defined by a probability density function (PDF), *p*_LoD_(*x*_lod_), in the domain of the normalized signal intensity, *x*_int_. The probability of positive agreement (i.e. the conditional positive agreement probability density function), is the corresponding cumulative distribution function (CDF) of the LoD PDF,

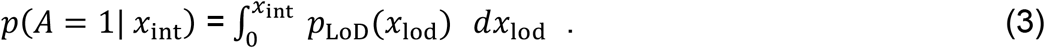

Correspondingly, the LoD PDF in the signal intensity domain is the derivative of the probability of positive agreement in the same domain. It summarizes the process of observation and assessment of the testing device by the observer, or the observer population.

### Probability of positive agreement across viral load related domains

To follow, we transform the probability of agreement in the signal intensity domain (expression 3), to the rest of the viral load related domains: recombinant protein concentration, viral concentration and qRT-PCR cycles. The defined random variables and their causal dependencies are described in the Bayesian network of figure 2. Let us consider, first, the case of the protein concentration domain. As per the intensity analysis described previously, we experimentally estimate a conditional probability for the signal intensity given the recombinant protein concentration, *p*(***x***_int_| ***x***_prot_). We use this information to propagate the probability of positive agreement from the signal intensity domain to the protein concentration domain. For this purpose, we apply the probability chain rule,

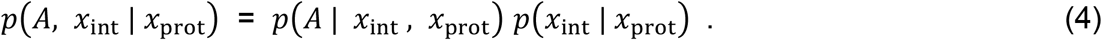

**Figure 2.**
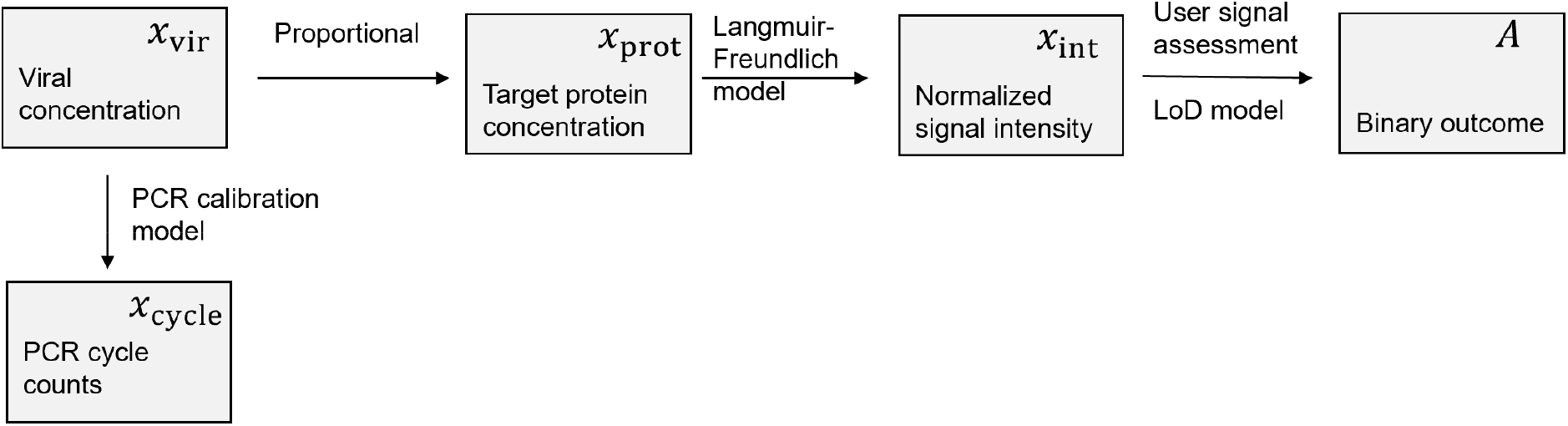
Bayesian network showing (boxes) the model random variables, (arrows) their causal relations and (annotations) relation models.

Integrating in ***x***_int_ domain, and taking into account that the observer assessment is only dependent on normalized signal intensity (see figure 2), *p*(*A* | *x*_int_, *x*_prot_) = *p*(*A* | *x*_int_),

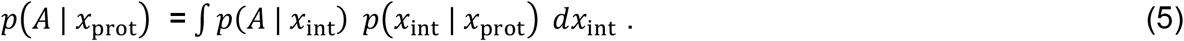

The functions within the integral involve the probability of positive agreement in the signal intensity domain (expression 3) and the PDF of signal intensity conditioned to the protein concentration. We perform the integral by Monte Carlo integration. Similarly, we transform the probability of agreement to the virus concentration domain and qRT-PCR cycles domain,

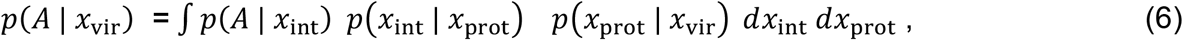

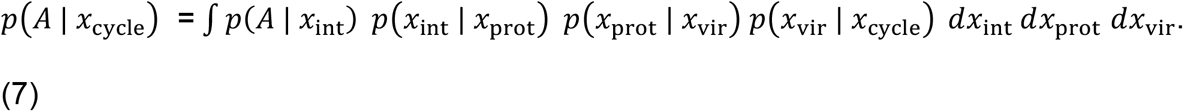

To model the probability of agreement in the domain of qRT-PCR cycles, as common for real-world testing, we solve equation 7 by Monte Carlo integration. We require for this purpose estimated models for the four conditional probabilities within the right-hand integrand, which involve the information represented in figures 1 and 2. In our implementation, we first integrate in the virus concentration domain, to have a relationship between the protein concentration and the qRT-PCR cycles, *p*(*x*_prot_ | *x*_cycle_). Thus,

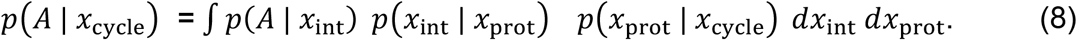

The conditional probability of positive agreement, *p*(*A* | *x*_cycle_), fully describes the test performance in the qRT-PCR cycle domain. For a given PDF of the sample PCR cycles distribution, the resulting sensitivity, *p*(*A*), is by integration,

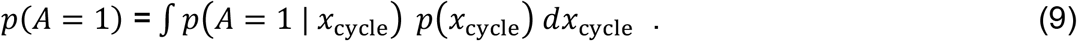

For a given collection of *N* real-world samples with PCR cycles, **x**_cycle_ = {*x*_l_, *x*_2_, … *x*_n_, …, *x*_N_ } the above integral is approximated by the average of the PPA function evaluated at the sample qRT-PCR cycles,

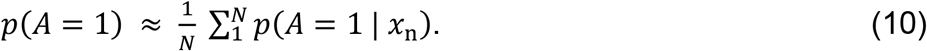

## Results

We illustrate the application of the described methodology with the characterization of the analyzed COVID-19 antigen test brand, and predicting the corresponding real-world probability of agreement conditioned to qRT-PCR data.

Figure 3 shows the signal intensity data corresponding to protein dilutions prepared in the laboratory for this AT, and the corresponding Langmuir-Freundlich regression model; from the analysis we model the conditional PDF, *p*(*x*_int_ | *x*_prot_). Figure 4 illustrates our modeled relationships across various viral load related variables, based on our experimental characterization of the antigen test and qRT-PCR calibration. Figure 4a shows the signal intensity analysis antigen test, based on serial dilutions of inactivated virus. The plot is similar to figure 3, which describes the signal response to protein dilutions. By combining the protein and virus curves for common signal intensity responses, we calibrate a linear model that describes the relationship between protein and inactivated virus concentration (figure 4b). The figure 4c shows the calibration of the qRT-PCR cycles (Ct) curve based on PCR analyzed inactivated virus dilution series. The qRT-PCR analysis of the dilution series was conducted by the same center used for self-testing of our ordinary (real-world) testing program. Figure 4d shows the qRT-PCR cycles and protein concentration relation, transformed from the viral concentration domain by the protein-virus relationship characterized in figure 4b. All the relationships shown in figure 4, are modeled as conditional PDFs; for illustration the plots show specific confidence limits.

**Figure 3.**
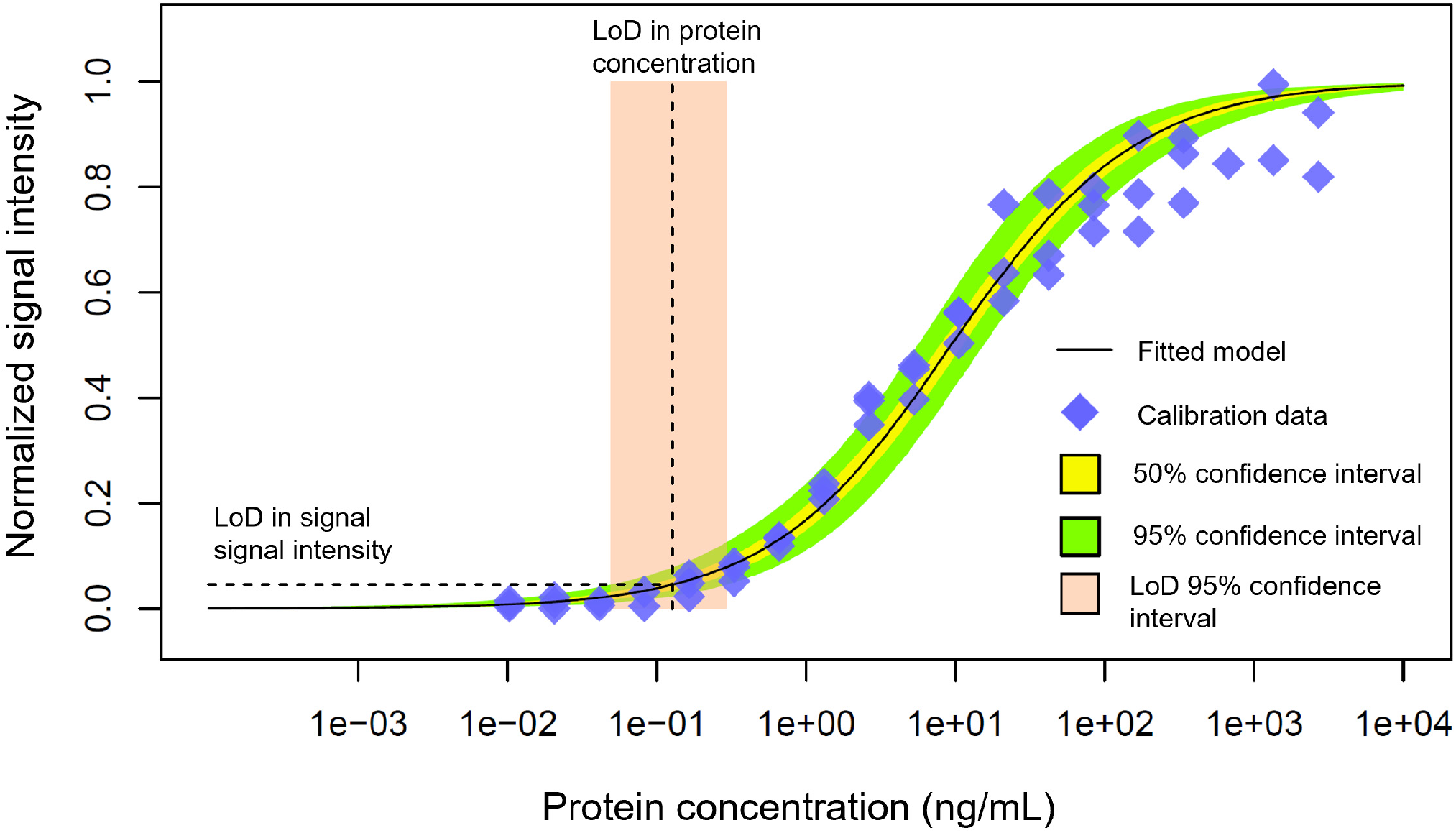
Normalized signal intensity data for three series of protein dilutions for the analyzed Covid-19 test brand, and the Langmuir-Freundlich model estimated to fit the data. The estimation also involves the uncertainties in the data fit, as shown by the confidence interval bands. The dashed line shows the projection to the protein concentration domain of an assumed LoD of 5% in the normalized signal intensity domain.

**Figure 4.**
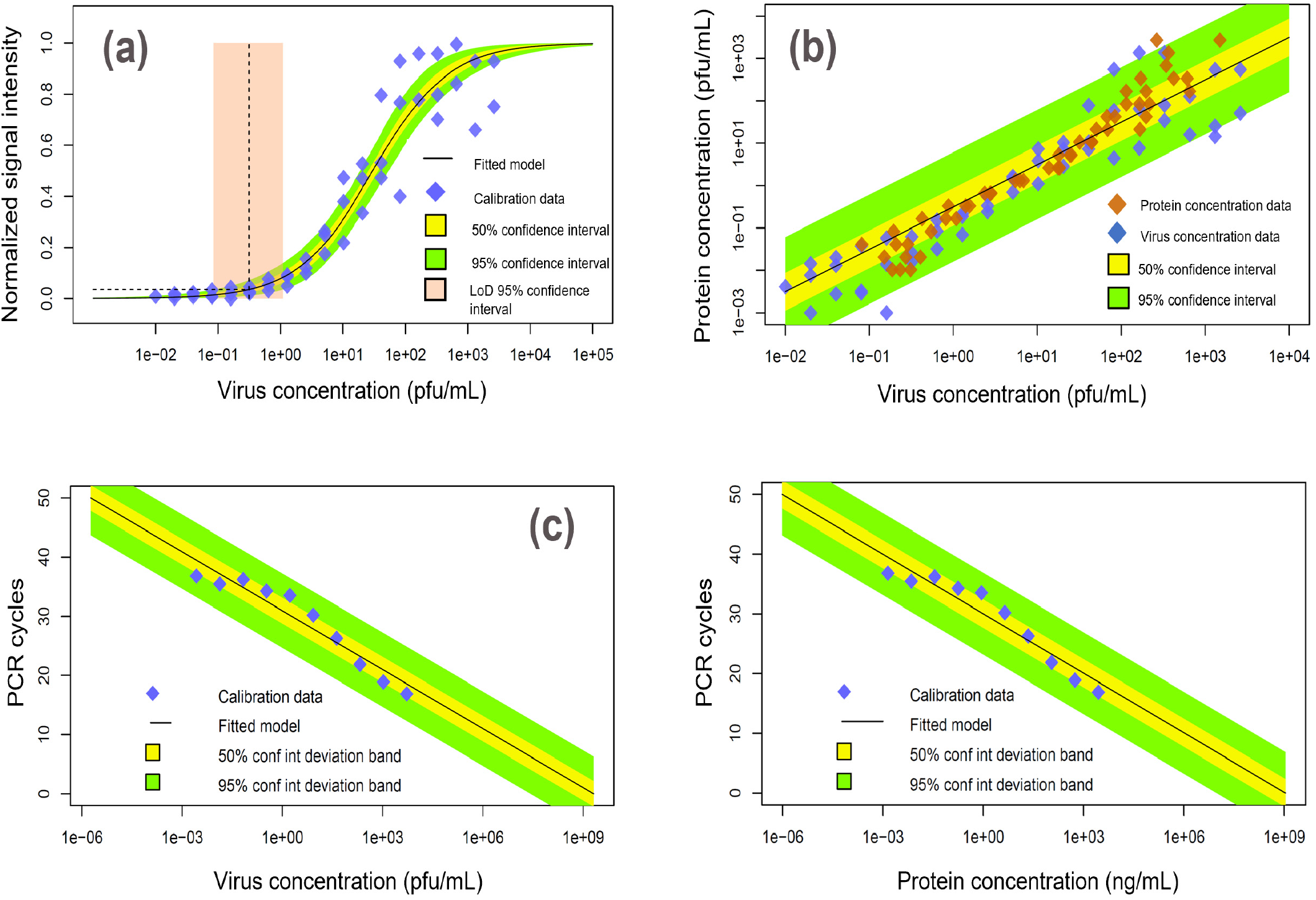
Characterization of the signal response with inactivated virus and domain related transformation functions calibrated from experimental data. (a) The normalized signal response analysis for serial dilutions of inactivated virus. (b) Protein and virus concentration linear relational model based on common signal intensity response of the devices, which allows transforming virus to protein concentration and vice-versa. (c) PCR cycle response calibration to inactivated virus dilution series. (d) qRT-PCR cycle response related to protein concentration, by combining transformations (b) and (c).

An additional component required by our predictive method is the LoD PDF of the observer’s population, *p*_LoD_(*x*_lod_), in the domain of the signal intensity. Figure 5a, shows the real-world binary assessment conditioned to the signal intensity (based on the Boston, Chelsea area AT results and photos uploaded by the participants), the corresponding PPA function of the signal intensity estimated by logistic regression, and the LoD PDF in the domain of the signal intensity. The former is the observed LoD in the domain of the signal intensity for the participant’s population. Although, we can characterize the LoD in the domain of the signal intensity based on the real-world naked-eye data of the participant’s population, it is interesting to compare this estimation to one based on easier to obtain naked-eye data. Figure 5b, shows naked-eye data, PPA function and LoD PDF, for our company’s personnel assessing the result on the antigen test activated by a series of recombinant protein dilutions (including zero concentration cases), after blinding any concentration labels. Regardless of the important differences in testing population and conditions, the probability functions (cumulative and density) are remarkably similar across figures 5a and 5b.

**Figure 5.**
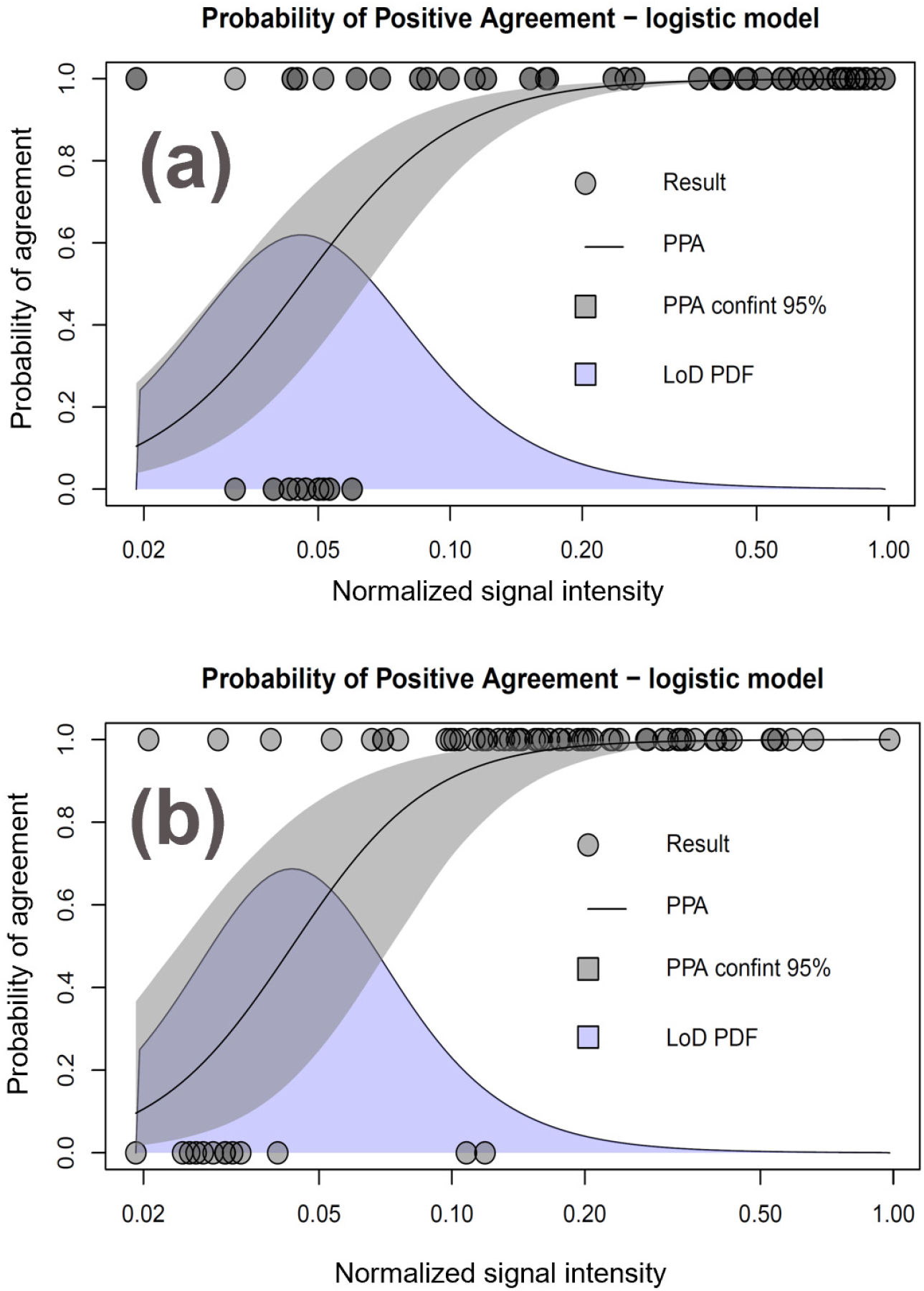
Probability of positive agreement and limit of detection (LoD) probability density for naked eye assessments of antigen tests in common cassette presentation made by two different groups of observers: a) company’s personnel observing devices activated by protein dilutions with concentration annotations blinded, and b) community participants from Boston’s Chelsea area observing their own self-tested results. In the former case the normalized intensity was calculated from laboratory photographs of the devices. In the latter case it was calculated from self-loaded participant mobile phone photographs uploaded in our reporting system.

With the analysis shown in figures 3-5 that model the input components of the predictive model, we perform our calculations of the PPA function of the qRT-PCR cycles and compare it with the *observed* PPA function (i.e. derived from the real-world data). Figure 6a shows the real-world data of binary results obtained with the antigen test (i.e. self-tested and logged data in Boston, Chelsea area project), and the corresponding observed PPA as function of the qRT-PCR cycles. The calculation involves the description of the model uncertainties, illustrated in the plot with the confidence intervals.

**Figure 6.**
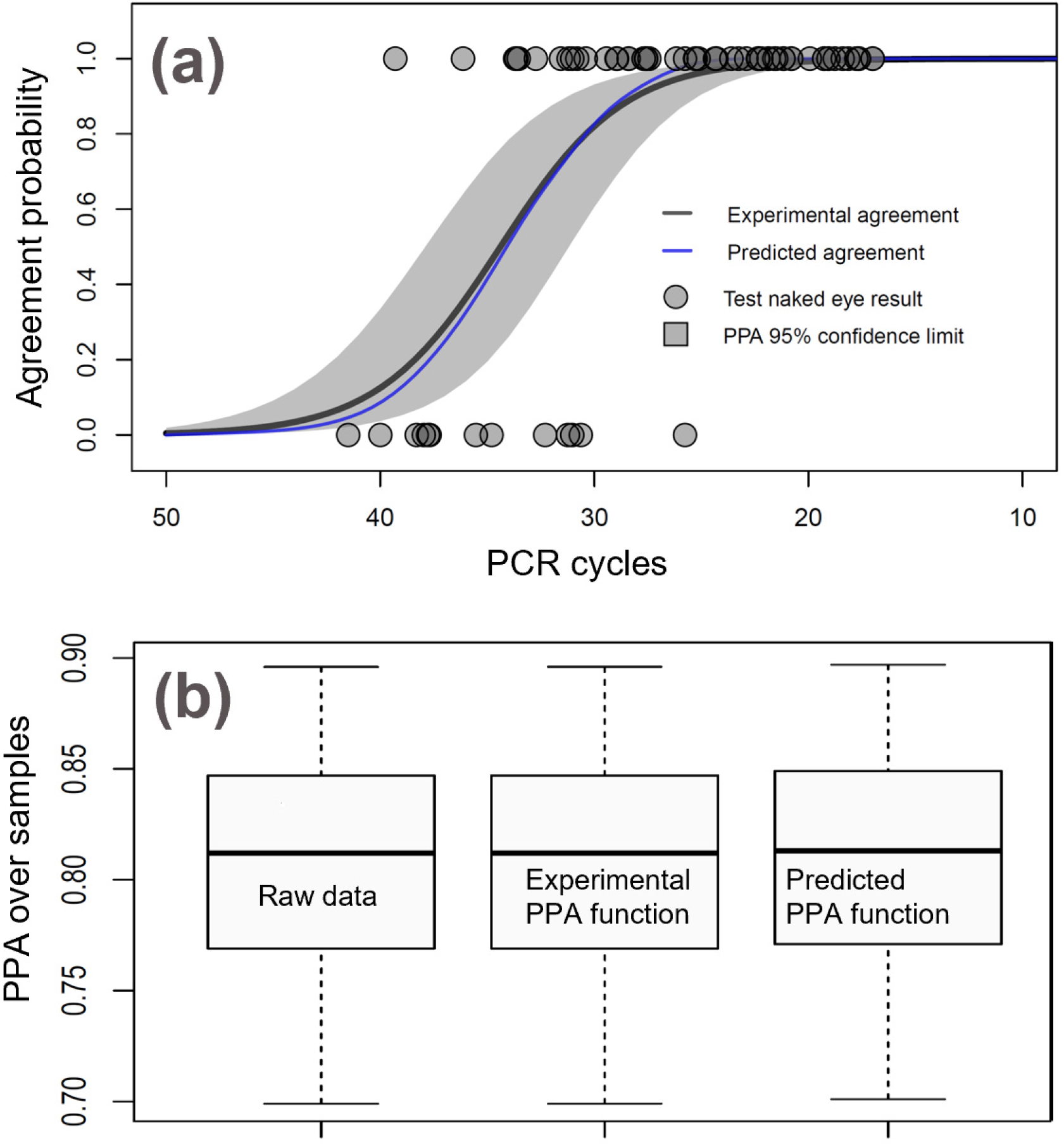
(a) Probability of agreement functions based on (black curve) experimental real-world data, and (blue curve) resulting from our predictive model. The experimental PPA function was obtained from a logistic regression analysis of the Chelsea project data for the test users; the gray area shows 95% confidence limits on the experimental PPA function. (b) Boxplots showing the PPA calculated over the real-world raw data, the observed PPA function and the predicted PPA function. Boxplot bounds are Clopper-Pearson confidence limits for percentiles (box) 50% and (segment) 95%.

Superimposed, we represent the predicted PPA function in the domain of the qRT-PCR, calculated by our method (expression 8). The plot clearly shows that the predicted and observed PPA functions are very close and within the figure confidence limits. Figure 6b shows a boxplot of the overall data PPA for samples collected in the real-world setting for the AT, and a comparison of the corresponding predicted real-world PPA according to expression 10. We can verify that both boxplots provide close results, within statistical significance.

## Discussion

The formulation for our predictive model and the application example shows that real-world behavior of antigen tests, can be reliably anticipated from quick laboratory testing and characterization of the population LoD for the devices used. First, the analysis of signal intensity and calibration of the qRT-PCR can be performed entirely under laboratory conditions; a fast process that precisely evaluates antigen test performance. Second, we have shown that the statistical characterization of the LoD of the observer’s population in the interpretation of common AT devices is feasible task. We also showed with experimental data that the LoD PDF remained very similar with two different observer population sub-groups and testing conditions. In principle, the LoD in the domain of the device signal intensity should be dependent on the observer’s visual capacity and device’s design (e.g. cassette, strip, card). That means that the characterization of the prior LoD PDF could be performed with statistics over the user population, with devices activated by protein, inactivated virus or any other safe recombinant reactions – it does not require clinically controlled experiments.

We think that these laboratory analyses and the predictive model methodology are useful information for regulatory evaluation of antigen tests. Better informed decision making may involve: (1) quick preliminary authorizations, to be followed with real-world data validation, (2) lab testing to validate real-world data provided by applicants, (3) considering regulations based entirely on the quantitative performance of the tests in the laboratory, and (4) easier post-approval quality control of the AT brands.

The accurate calibration of the relational components (figure 2) is fundamental for the accuracy of the model. We followed with care dilution factors along the process of sample preparation for analysis. Also, we verified that thermal inactivation of the virus, as employed in the lab serial dilutions, involves a degradation of the capsid protein of approximately 2x, which we accounted for as a dilution factor. We also used the same inactivated virus stock for the signal intensity and PCR cycle count calibration.

The purpose of the present manuscript is the description of a method for quantification and prediction of antigen test performance. Further work is in progress, searching for expanding our data for comparative analysis of performance prediction across several test brands, comparative calibration of the qRT-PCR cycles across service providers, and more data on the characterization of the signal intensity LoD for common antigen test

## Conclusions

An accurate description of the antigen test signal intensity response conditioned by variables related to viral load, such as concentration of recombinant protein and concentration of inactivated virus, can be stablished under laboratory conditions. These evaluations involve image processing of photographs and human naked-eye assessments on elaborated dilution series. Modeling the performance on real testing conditions involves integrating the mentioned laboratory evaluation with information on the ability of the observer population in recognizing the device visual response. This ability can be described by the LoD PDF of the observer population in the domain of the test signal intensity. Finally, we describe the test performance with the PPA *function* of the qRT-PCR cycles, instead of the common plain sample PPA (i.e. sensitivity). The presented methodology has promising applications for regulatory science regarding the evaluation of antigen tests, as it involves a quick appraisal of the real-world test performance.

## Data Availability

All data is available at the Rapid Acceleration of Diagnostics - Underserved Populations (RADxUP, a program funded by the National Institutes of Health (NIH), under the repository conditions.

https://idx20.us

## Acknowledgments

We gratefully acknowledge the collaboration of the Center for Complex Intervention, the Chelsea Housing Authority, the Chelsea City Health Department, and the study participants in this research. We also extend our sincere thanks to the Reagan-Udall Foundation for their generous support through Grant #02282022 RUF, which facilitated the development of this study.

## Notes

**Funding:** This research was supported by Reagan-Udall Foundation through Grant #02282022 RUF, awarded to IDX20.

**Declaration of interests:** Irene Bosch is a founder of IDX20, a company affiliated with this study. Miguel Bosch is a founder of Info Analytics Innovations, a company also affiliated with this study. The authors declare no additional conflicts of interest.

### Competing Interest Statement

Irene Bosch is the founder and owner of IDX20, a company affiliated with this study. Miguel Bosch is the founder and owner of Info Analytics Innovations, a company also affiliated with this study. The authors declare no additional conflicts of interest.

### Funding Statement

This research was supported by Reagan-Udall Foundation through Grant #02282022 RUF, awarded to IDX20.

### Author Declarations

Advarra gave ethical approval for this work

